# A Generalized Signal Model for Dual-Module Velocity-Selective Arterial Spin Labeling

**DOI:** 10.1101/2024.05.15.24307357

**Authors:** Thomas T. Liu, Conan Chen, Jia Guo, Eric C. Wong, Divya S. Bolar

**Author notes:** **Correspondence** Thomas T. Liu, Center for Functional MRI, University of California San Diego, 9500 Gilman Drive MC 0677, La Jolla, CA 92093.

## Abstract

**Purpose:** To develop a generalized signal model for dual-module velocity-selective arterial spin labeling (dm-VSASL) that can integrate arbitrary saturation and inversion profiles.

**Theory and Methods:** A recently developed mathematical framework for single-module VSASL is extended to address the increased complexity of dm-VSASL and to model the use of realistic velocity-selective profiles in the label-control and vascular crushing modules. Expressions for magnetization difference, arterial delivery functions, labeling efficiency, and cerebral blood flow (CBF) estimation error are presented. Sources of error are examined and timing requirements to minimize quantification errors are derived.

**Results:** For ideal velocity-selective profiles, the predicted signals match those of prior work. With realistic profiles, a CBF-dependent estimation error can occur when velocity-selective inversion (VSI) is used for the labeling modules and velocity-selective saturation (VSS) is used for the vascular crushing module. The error reflects a mismatch between the leading and trailing edges of the delivery function for the second bolus and can be minimized by choosing a nominal labeling cutoff velocity that is lower than the nominal saturation cutoff velocity. In the presence of *B*_0_ and *B*_1_ inhomogeneities, the labeling efficiency of dual-module VSI is more attenuated than that of dual-module VSS.

**Conclusion:** The proposed signal model will enable researchers to more accurately assess and compare the performance of realistic dm-VSASL implementations and improve the quantification of dm-VSASL CBF measures.

## 1 INTRODUCTION

Dual-module velocity-selective arterial spin labeling (dm-VSASL) has been shown to offer improvements in robustness and SNR efficiency as compared to single-module VSASL^1,2,3,4,5^. The primary gain in SNR efficiency has been shown to reflect the creation of a larger magnetization difference between the control and label conditions through re-labeling of spins by the second module^1,2,3^. When using modules that invert static tissue, dm-VSASL offers inherent background suppression that can further improve performance and robustness^2,6^. Although typically negligible, there can also be an SNR efficiency gain due to an increase in bolus duration that reflects labeling by the second module of arterial spins that have entered the coverage of the RF transmit coil after the first module^1,2,3^.

Prior dm-VSASL signal models have assumed ideal saturation and inversion profiles and are not easily extended to handle realistic profiles. Here we build upon a recently introduced mathematical model for single-module VSASL^7^ to develop a generalized signal model for dm-VSASL that can readily integrate realistic saturation and inversion profiles and thus enable more accurate quantification of cerebral blood flow (CBF).

As the Theory section is rather involved, we start with a guide to the theoretical subsections. In Section 2.1, we review the basic structure and timing of a typical dm-VSASL scan and present the signal model. Then, in Section 2.2, we adopt the concept of passband and saturation reference functions introduced in Ref. [7] to rewrite the signal model in a form that makes explicit the dependence on the amplitudes and shapes of the label-control and saturation functions. In Section 2.3 we demonstrate the application of the framework with ideal rect profiles and compare the resulting expressions with those from prior work^2^. We present expressions for CBF quantification error and labeling efficiency in Sections 2.4 and 2.5, respectively, and then go on to show the dependence of the CBF quantification error on effective bolus width errors in Section 2.6 and present conditions for minimizing the errors in Section 2.7. Finally, we briefly consider cerebral blood volume effects in Section 2.8. Analyses using realistic Bloch-simulated profiles and taking into account CBF levels, varying timing parameters, and *B*_0_ and *B*_1_ inhomogeneities are described in the Methods and Results sections, and areas for future work are addressed in the Discussion.

## 2 THEORY

### 2.1 Signal Model

We consider the dual-module VSASL diagram shown in Figure 1, with two label-control modules (LCM1 and LCM2), a vascular crushing module (VCM), and timing parameters *T*_sat_, *τ*_1_, *τ*_2_, and PLD. To simplify the presentation, we assume that LCM1, LCM2, and the VCM are applied instantaneously at *t* = 0, *τ*_1_, and *τ*_1_ + *τ*_2_, respectively, such that TI = *τ*_1_ + *τ*_2_ + PLD. The labeling and control profiles are *l*_1_(*v*) and *c*_1_(*v*), respectively, for LCM1, and *l*_2_(*v*) and *c*_2_(*v*) for LCM2, where *v* denotes velocity. The labeling profiles can represent any velocity-selective implementation, including velocity-selective inversion (VSI), velocity-selective saturation (VSS), or velocity-selective saturation with static spins inverted (VSSI)^2^. The VCM saturation response is represented by *s*(*v*).

**FIGURE 1.**
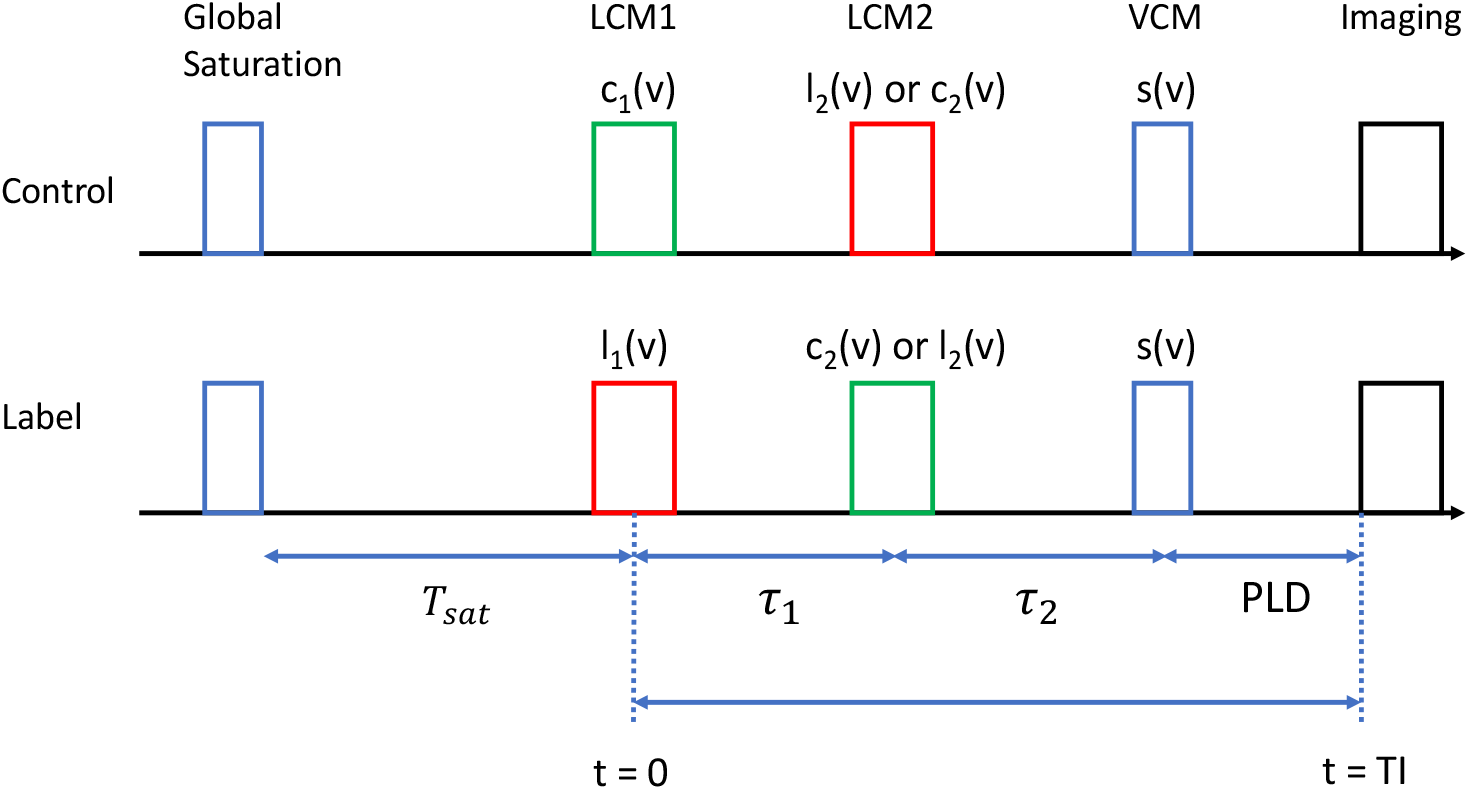
Dual-module VSASL timing diagram. For dmVSI and mixed variants (e.g. VSSI+VSS or VSI+VSS), LCM2 uses *l*_2_(*v*) and *c*_2_(*v*) in the control and label conditions, respectively. For dmVSS, LCM2 uses *c*_2_(*v*) and *l*_2_(*v*) in the control and label conditions, respectively.

We focus here on dual-module VSI (dmVSI) and dual-module VSS (dmVSS), as these are currently the most widely used dm-VSASL implementations. In LCM1, *c*_1_(*v*) and *l*_1_(*v*) are applied during the control and label conditions, respectively. For dmVSI, LCM2 uses *l*_2_(*v*) and *c*_2_(*v*) in the control and label conditions, respectively, whereas for dmVSS, *c*_2_(*v*) and *l*_2_(*v*) are applied in the control and label conditions, respectively. The expressions presented below for dmVSI also apply to other combinations^2^ (which we will refer to as mixed variants), such as VSSI+VSS and VSI+VSS, that swap the label and control responses between LCM1 and LCM2. A glossary of key functions and variables is provided in Table 1.

**TABLE 1.** Glossary of key functions and variables. To simplify the presentation we have omitted subscripts that can be used to indicate different values of the functions and variables for LCM1 and LCM2, e.g. subscripts can be added to *l*(*v*) to denote *l*_1_(*v*) and *l*_2_(*v*).

| Symbol | Description |
| --- | --- |
| $T_{sat}$ | Saturation Time |
| $\tau_1, \tau_2$ | Bolus timing parameters |
| PLD | Post-labeling delay |
| TI | Inflow time |
| VSS | Velocity selection saturation |
| VSI | Velocity selection inversion |
| $l(v)$ | Labeling profile |
| $c(v)$ | Control profile |
| $s(v)$ | Saturation function: $s(v) = S_0 \cdot s_{ref}(v)$ |
| $p(v)$ | Passband function: $p(v) = P_a \cdot p_{ref}(v)$ |
| $p_{ref}(v)$ | Reference Passband function with $p_{ref}(\infty) = 1$ |
| $s_{ref}(v)$ | Reference Saturation function with $s_{ref}(0) = 1$ |
| $s_{ref,E}(v)$ | Effective saturation function: $s_{ref,E}(v) = 1 - p_{ref}(v)$ |
| $P_a$ | Steady state value of $p(v)$; with ideal value $P_{a*} = 1$ for VSS and 2 for VSI |
| $S_0$ | Initial value $s(0)$; with ideal value of 1.0 |
| $\Delta M_{z,1}(t), \Delta M_{z,2}(t)$ | Cumulative magnetization differences |
| $L_1(t), L_2(t)$ | Longitudinal relaxation weighting functions |
| $d_{a,1}(t), d_{a,2}(t)$ | Arterial delivery functions |
| $\tau_{1,eff}, \tau_{2,eff}, \tau_{[1,2],eff}$ | Effective bolus widths; with ideal values of $\tau_1, \tau_2$, and $\tau_1 + \tau_2$ |
| $A_{1,eff}, A_{2,eff}$ | Effective bolus areas |
| $\kappa$ | Scaling term to denote common attenuation of $l(v)$ and $c(v)$ at $v = 0$ |
| $\beta$ | Additional scaling term for $p(v)$ |
| $\alpha$ | Labeling efficiency |
| $v_l$ | Label/Control module (LCM) nominal cutoff velocity |
| $v_c$ | Vascular crushing module (VCM) cutoff velocity |
| $v_a(t)$ | Arterial delivery velocity function $v_a(t, v_b)$ with implicit argument $v_b$ |
| $v_b$ | Boundary velocity |
| $v_{cap}$ | Mean capillary velocity |
| $Q_0$ | Cerebral blood flow (CBF) |
| $\Lambda(M_b, t)$ | Longitudinal recovery function |
| $\Delta t_l, \Delta t_c$ | LCM (subscript l) and VCM (subscript c) transit delays; defined as the times for arterial blood to decelerate from $v_l$ and $v_c$, respectively, to the boundary velocity $v_b$ |

Building on the framework presented in Ref. [7], it is shown in the Appendix that the cumulative magnetization difference (control-label) as a function of time for *t* > *τ*_1_ + *τ*_2_ is given by Δ*M*_z_(*t*) = Δ*M*_z,1_(*t*) + Δ*M*_z,2_(*t*) where

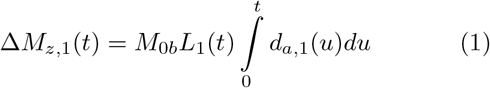

and

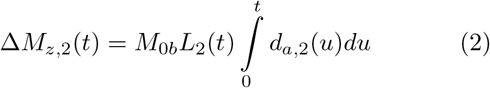

with longitudinal relaxation weighting functions

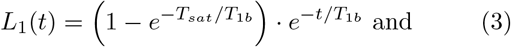

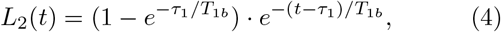

and *M*_0b_ and *T*_1b_ denoting the equilibrium magnetization and longitudinal relaxation time constant, respectively, of arterial blood. Equivalent arterial delivery functions are defined as

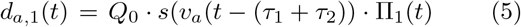

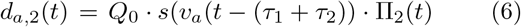

where

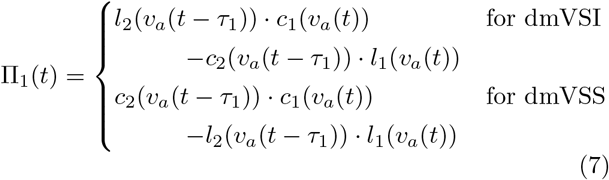

and

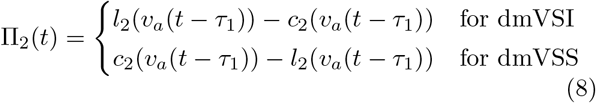

with *Q*_0_ denoting CBF. The function *v*_a_(*t*) denotes the initial velocity (at *t* = 0) of arterial blood that will decelerate to a boundary velocity *v*_b_ at a later time *t*, which was previously denoted as *v*_a_(*t, v*_b_) in Ref. [7]. To simplify the presentation, we have made the dependence on *v*_b_ implicit here, such that *v*_a_(*t*) = *v*_a_(*t, v*_b_).

With these definitions, *d*_a,1_(*t*) and *d*_a,2_(*t*) represent the arterial blood flow that arrives at time *t* to the boundary velocity *v*_b_, where *d*_a,1_(*t*) reflects the blood delivered in the bolus created by LCM1, LCM2, and the VCM and *d*_a,2_(*t*) reflects the blood delivered in the bolus created by LCM2 and the VCM. We use the term *equivalent* arterial delivery function to reflect the fact that blood that has already been delivered at some time *t* can be affected by pulses applied at a later time^7^, and the effect of the “future” pulses is included in the definition of the delivery function. For example, the value of *d*_a,1_(*t*) at some time 0 ≤ *t* < *τ*_1_ + *τ*_2_ reflects the application of the VCM at a future time *t* = *τ*_1_ + *τ*_2_.

### 2.2 Passband and Saturation Reference Functions

Following the approach described in Ref. [7], we define the passband function for the *i*th LCM module as 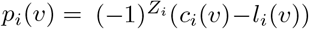 where *Z*_i_ is 1 for modules that invert static spins and 0 otherwise. It is useful to express the passband function as the product *p*_i_(*v*) = *P*_a,i_ · *p*_ref,i_(*v*) of an amplitude term *P*_a,i_ that denotes the steady-state value of the passband function and a reference passband function *p*_ref,i_(*v*) that starts at zero and approaches 1.0 for velocities above a labeling cut-off velocity *v*_l,i_. Similarly, it is useful to express the VCM saturation function as the product *s*(*v*) = *S*_0_ · *s*_ref_(*v*) of an amplitude term *S*_0_ = *s*(0) and a reference saturation function *s*_ref_(*v*) that starts at *s*_ref_(0) = 1.0 and approaches zero for velocities above a VCM cutoff velocity *v*_c_.

To facilitate comparison with prior work^2^, it is helpful to express the passband amplitude as the product *P*_a,i_ = *P*_a⋆,i_ · *κ*_i_ · *β*_i_ where *P*_a⋆,i_ denotes the ideal steadystate value (1.0 and 2.0 for VSS and VSI, respectively), *κ*_i_ = |*l*_i_(0)| = |*c*_i_(0)| ≤ 1.0 represents the absolute value of the control and label functions at *v* = 0 and reflects factors (such as *T*_1_, *T*_2_, and *B*_1_ effects) that equally affect both the label and control, and *β*_i_ ≥ 0 reflects remaining factors that are not accounted for by *κ*_i_, such as the steady-state value of the VSI labeling profile^7^.

With these definitions, we can immediately write

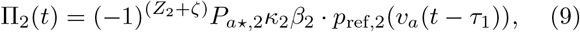

where *ζ* = 0 for dmVSS and *ζ* = 1 for dmVSI and mixed variants. With the additional assumption^2^ that *c*_i_(*v*) = *c*_i_(0), we can write for dmVSI (and mixed variants)

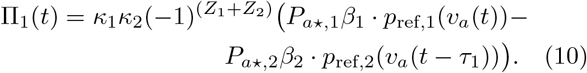

For a typical dmVSI implementation where the LCM modules use the same train of pulses this can be written as

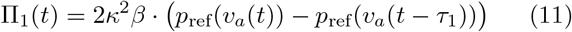

where *p*_ref_(*v*) is an index-independent reference passband function and *κ* and *β* are index-independent scaling terms.

For dmVSS, we have

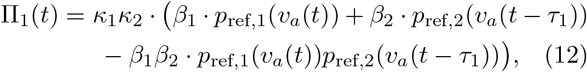

which for a typical implementation with identical LCM modules reduces to

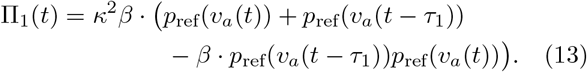

In practice, *p*_ref_(*v*_a_(*t*−*τ*_1_))*p*_ref_(*v*_a_(*t*)) ≈ *p*_ref_(*v*_a_(*t*−*τ*_1_)) for *τ*_1_ > Δ*t*_l_, where Δ*t*_l_ is the LCM transit delay^7^, resulting in

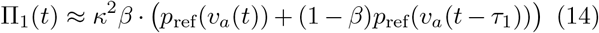

### 2.3 Ideal Profiles

We first consider delivery functions obtained with ideal passband *p*_ref_(*v*) = 1 − rect(*v/*(2*v*_l_)) and saturation *s*_ref_(*v*) = rect(*v/*(2*v*_c_)) functions, where *v*_l_ and *v*_c_ are the labeling and saturation cutoff velocities^7^, respectively, and assumed to be equal unless otherwise noted. Examples of *d*_a,1_(*t*) and *d*_a,2_(*t*) are shown in Figs. 2A and 3A for dmVSI and dmVSS, respectively, with *Q*_0_ = 1, *τ*_1_ = 1.5 s, *τ*_2_ = 0.5 s, *v*_b_ = 0.1 cm/s, *v*_l_ = *v*_c_ = 2 cm/s, and *κ* = *β* = *S*_0_ = 1.0. Additional examples with realistic Bloch-simulated profiles are discussed in the Results section.

**FIGURE 2.**
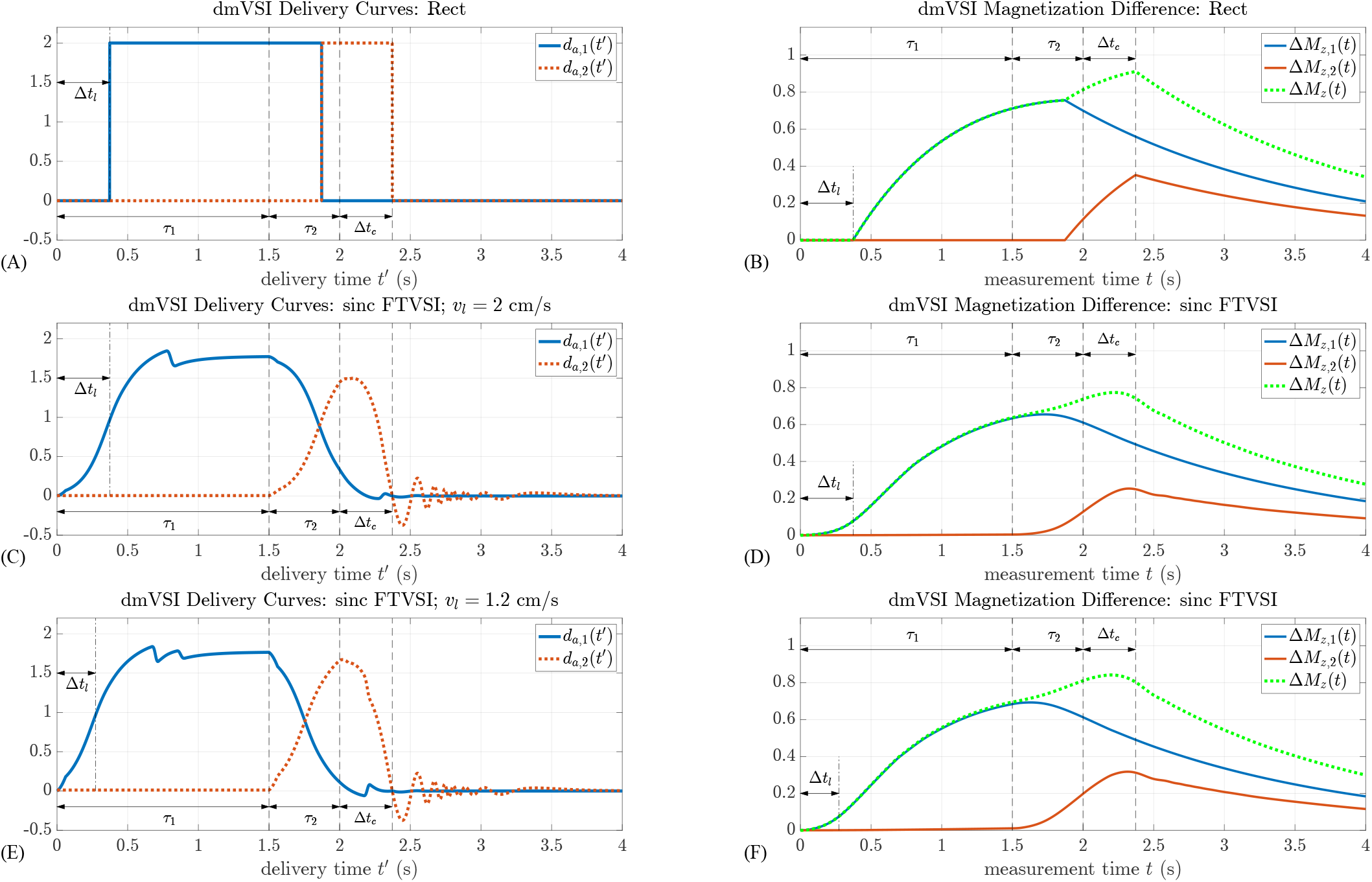
dmVSI equivalent arterial delivery functions. (A, C, E) and cumulative magnetization differences (B, D, F) for ideal rect profile (top row) and Bloch-simulated sinc FTVSI nominal profiles (second and third rows) with *Q*_0_ = 1, *τ*_1_ = 1.5 s, *τ*_2_ = 0.5 s, *v*_*b*_ = 0.1 cm/s, *v*_*c*_ = 2 cm/s, *v*_*l*_ = 2 cm/s (top 2 rows) or *v*_*l*_ = 1.2 cm/s (bottom row), and Δ*t*_*l*_ and Δ*t*_*c*_ denoting the LCM and VCM transit delays^7^. For the top two rows, these transit delays are equal and indicate the time needed for arterial blood to decelerate from 2 cm/s to 0.1 cm/s. For the bottom row, Δ*t*_*l*_ < Δ*t*_*c*_ since *v*_*l*_ < *v*_*c*_, with Δ*t*_*l*_ indicating the time for arterial blood to decelerate from 1.2 cm/s to 0.1 cm/s. Note that area under the curve of *d*_*a*,2_(*t*) for *v*_*l*_ = 1.2 cm/s (panel E) is greater than that for *v*_*l*_ = 2.0 cm/s (panel C), reflecting the better matching of the leading and trailing edges in the delivery function for the second bolus. This results in greater amplitudes in the Δ*M*_*z*,2_(*t*) and Δ*M*_*z*_(*t*) curves for *v*_*l*_ = 1.2 cm/s. The delivery time *t*^*′*^ and measurement time *t* variables are described in the Appendix. Curves for Δ*M*_*z*,1_(*t*), Δ*M*_*z*,2_(*t*), and Δ*M*_*z*_ (*t*) are depicted for *t* ≥ 0, but reflect what is actually measured only for the period after application of the VCM, i.e. *t* ≥ *τ*_1_ + *τ*_2_. For *t* < *τ*_1_ + *τ*_2_, they reflect the “virtual” magnetization difference that would be measured if the effects of future events (e.g. LCM2 and VCM) could affect prior periods.

**FIGURE 3.**
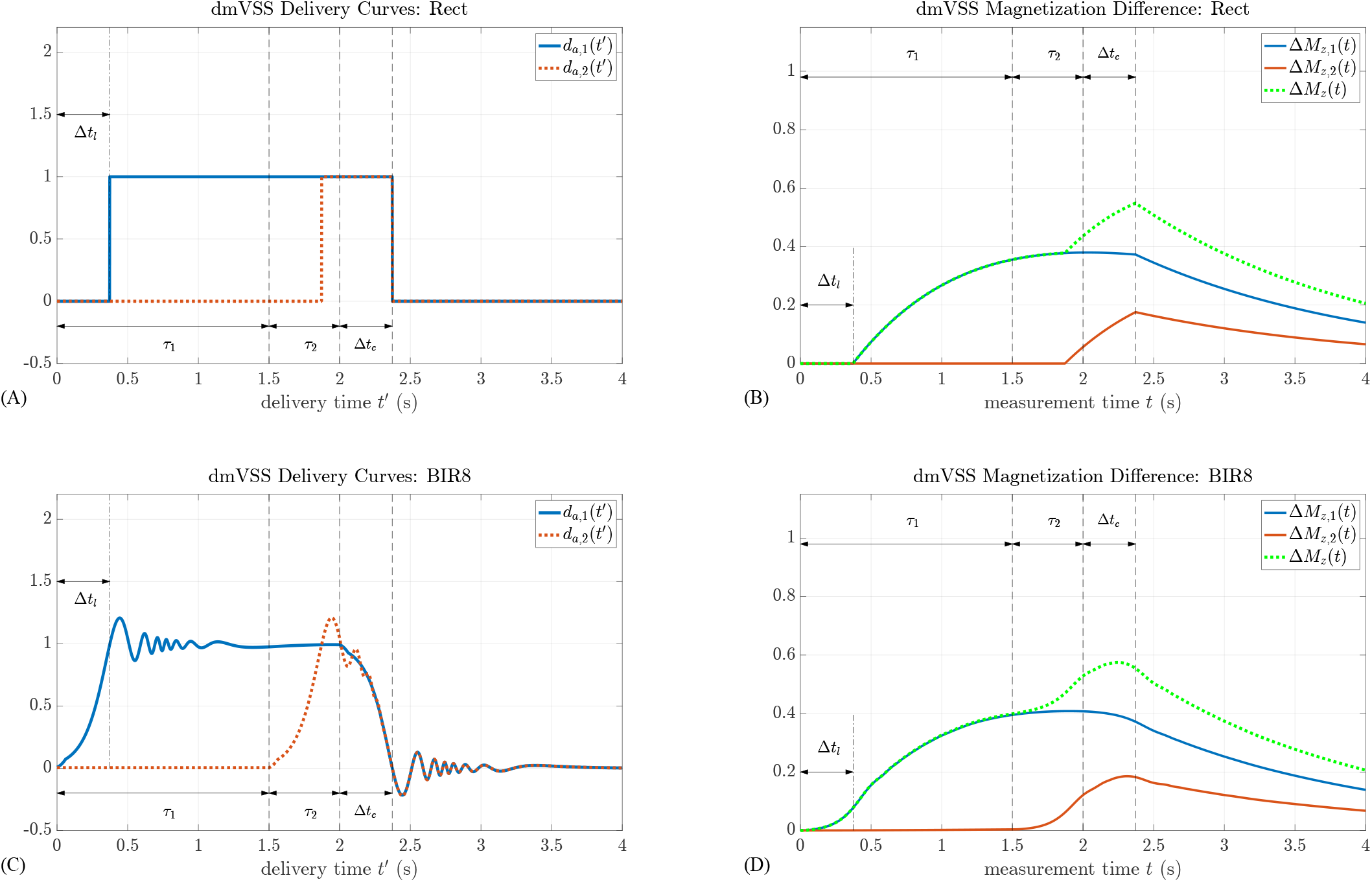
dmVSS equivalent arterial delivery functions. (A, C) and cumulative magnetization differences (B,D) for ideal rect profile (top row) and Bloch-simulated BIR8 nominal profile (bottom row) with *Q*_0_ = 1, *τ*_1_ = 1.5 s, *τ*_2_ = 0.5 s, *v*_*b*_ = 0.1 cm/s, *v*_*l*_ = *v*_*c*_ = 2 cm/s, and Δ*t*_*l*_ and Δ*t*_*c*_ denoting the labeling and VCM transit delays^7^, which are equal for this example and indicate the time needed for arterial blood to decelerate from 2 cm/s to 0.1 cm/s. The delivery time *t*^*′*^ and measurement time *t* variables are described in the Appendix. Curves for Δ*M*_*z*,1_(*t*), Δ*M*_*z*,2_(*t*), and Δ*M*_*z*_ (*t*) are depicted for *t* ≥ 0, but reflect what is actually measured only for the period after application of the VCM, i.e. *t* ≥ *τ*_1_ + *τ*_2_. For *t* < *τ*_1_ + *τ*_2_, they reflect the “virtual” magnetization difference that would be measured if the effects of future events (e.g. LCM2 and VCM) could affect prior periods.

For dmVSI, *d*_a,1_(*t*) and *d*_a,2_(*t*) in Fig. 2A correspond to ideal boluses with amplitudes of 2*Q*_0_ spanning the intervals [Δ*t*_l_, *τ*_1_ + Δ*t*_l_] and [*τ*_1_ + Δ*t*_l_, *τ*_1_ + *τ*_2_ + Δ*t*_c_], respectively, with corresponding bolus areas of 2*Q*_0_*τ*_1_ and 2*Q*_0_*τ*_2_, where Δ*t*_c_ is the VCM transit delay^7^ and Δ*t*_l_ and Δ*t*_c_ are equal for this example and represent the time for arterial blood to decelerate from *v*_l_ = *v*_c_ = 2 cm/s to 0.1 cm/s. The corresponding cumulative magnetization differences, Δ*M*_z,1_(*t*) and Δ*M*_z,2_(*t*), and the sum Δ*M*_z_(*t*) are shown in Fig. 2B, where all spins are delivered by *t* = *τ*_1_ + *τ*_2_ +PLD as the PLD is set equal to Δ*t*_c_. The selection of PLD is discussed in detail in Section 2.7. For general values of *S*_0_, *κ*, and *β*, the bolus areas of *d*_a,1_(*t*) and *d*_a,2_(*t*) are 2*Q*_0_*S*_0_*κ*^2^*βτ*_1_ and 2*Q*_0_*S*_0_*κβτ*_2_. If 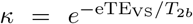 (where eTE_VS_ and *T*_2b_ denote the effective echo time and *T*_2_ of arterial blood), then the associated expressions for Δ*M*_z,1_(*t*) and Δ*M*_z,2_(*t*) match those for dmVSI groups 1 and 2, respectively, in Ref. [2]. For dmVSS, *d*_a,1_(*t*) and *d*_a,2_(*t*) in Fig. 3A correspond to ideal boluses with amplitudes of *Q*_0_ spanning the intervals [Δ*t*_l_, *τ*_1_ + *τ*_2_ + Δ*t*_l_] and [*τ*_1_ + Δ*t*_l_, *τ*_1_ + *τ*_2_ + Δ*t*_c_], respectively, with corresponding bolus areas of *Q*_0_(*τ*_1_ + *τ*_2_) and *Q*_0_*τ*_2_. The corresponding cumulative magnetization differences, Δ*M*_z,1_(*t*) and Δ*M*_z,2_(*t*), and the sum Δ*M*_z_(*t*) are shown in Fig. 3B. For general values of *S*_0_, *κ*, and *β*, the bolus areas of *d*_a,1_(*t*) and *d*_a,2_(*t*) are *Q*_0_*S*_0_*κ*^2^*β*(*τ*_1_ + *τ*_2_ + (1 − *β*)*τ*_2_) and *Q*_0_*S*_0_*κβτ*_2_, respectively. Comparing to Ref. [2], it can be shown that the associated expression for Δ*M*_z,2_(*t*) matches the first two terms of the dmVSS group 2 expression in the prior work, while the expression for Δ*M*_z,1_(*t*) contains a term that matches that of group 1 in the interval [Δ*t*_l_, *τ*_1_ + Δ*t*_l_] and another term that nearly matches the prior work’s remaining dmVSS group 2 terms in the interval [*τ*_1_ + Δ*t*_l_, *τ*_1_ + *τ*_2_ + Δ*t*_c_]. The discrepancy stems from a minor error in the prior derivation^2^, which is corrected in the current work.

### 2.4 CBF Quantification

An unbiased estimate of *Q*_0_ based on a measurement of the magnetization difference at *t* = TI can be written as

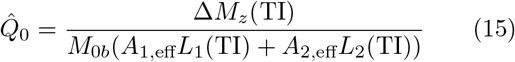

where

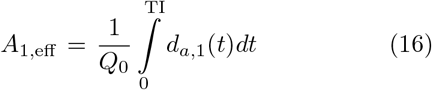

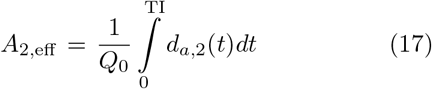

denote effective areas. If the estimate uses values 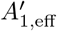 and 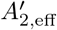 that differ from the actual values *A*_1,eff_ and *A*_2,eff_, then the resulting estimate 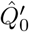 is biased. The fractional error of the biased estimate can be written as

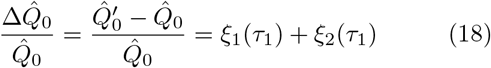

where

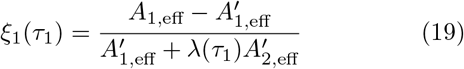

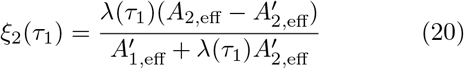

are the error components for the first and second boluses and

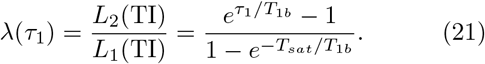

### 2.5 Labeling Efficiency

In the VSASL white paper^3^ (n.b. erratum), the difference signal for single-module VSASL has the form

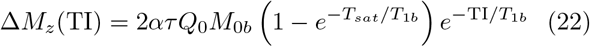

where *α* and *τ* were defined as the single-module VSASL labeling efficiency and bolus width, respectively. Comparing this to Eqn. 15, setting *τ* = *τ*_1_ + *τ*_2_, and equating like terms leads to

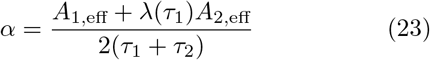

as the expression for dm-VSASL labeling efficiency.

For dmVSI with ideal rect profiles,

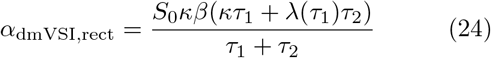

and for dmVSS with ideal rect profiles,

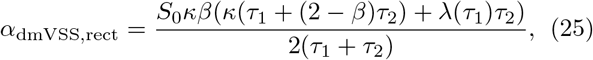

where we have assumed that the timing requirements specified below in Section 2.7 are satisfied. There do not appear to be analytical expressions for finding the maxima of *α*_dmVSI,rect_ and *α*_dmVSS,rect_, but they are straightforward to evaluate and optimize numerically.

Examples of ideal rect labeling efficiency versus *τ*_1_ are shown by the dashed green lines in Figs. 4A and 5A assuming *S*_0_ = *κ* = 1.0, *τ*_1_ + *τ*_2_ = 2.0 s, *T*_sat_ = 2.5 s, *T*_1b_ = 1.66 s, and *β* = 0.88 for dmVSI and *β* = 1.0 for dmVSS, where the dmVSI *β* = *P*_a_*/*2 value corresponds to the amplitude *P*_a_ = 1.76 of the sinc Fourier Transform VSI (FTVSI) passband function first examined in Ref. [7] and used later in this paper. Note that since *α* is referenced to the performance of single-module VSASL (with a maximum value of 1.0 for ideal VSI), the additional labeling of spins provided by dm-VSASL^1,2^can result in *α* > 1.0 under ideal conditions, as shown in Fig. 4A.

**FIGURE 4.**
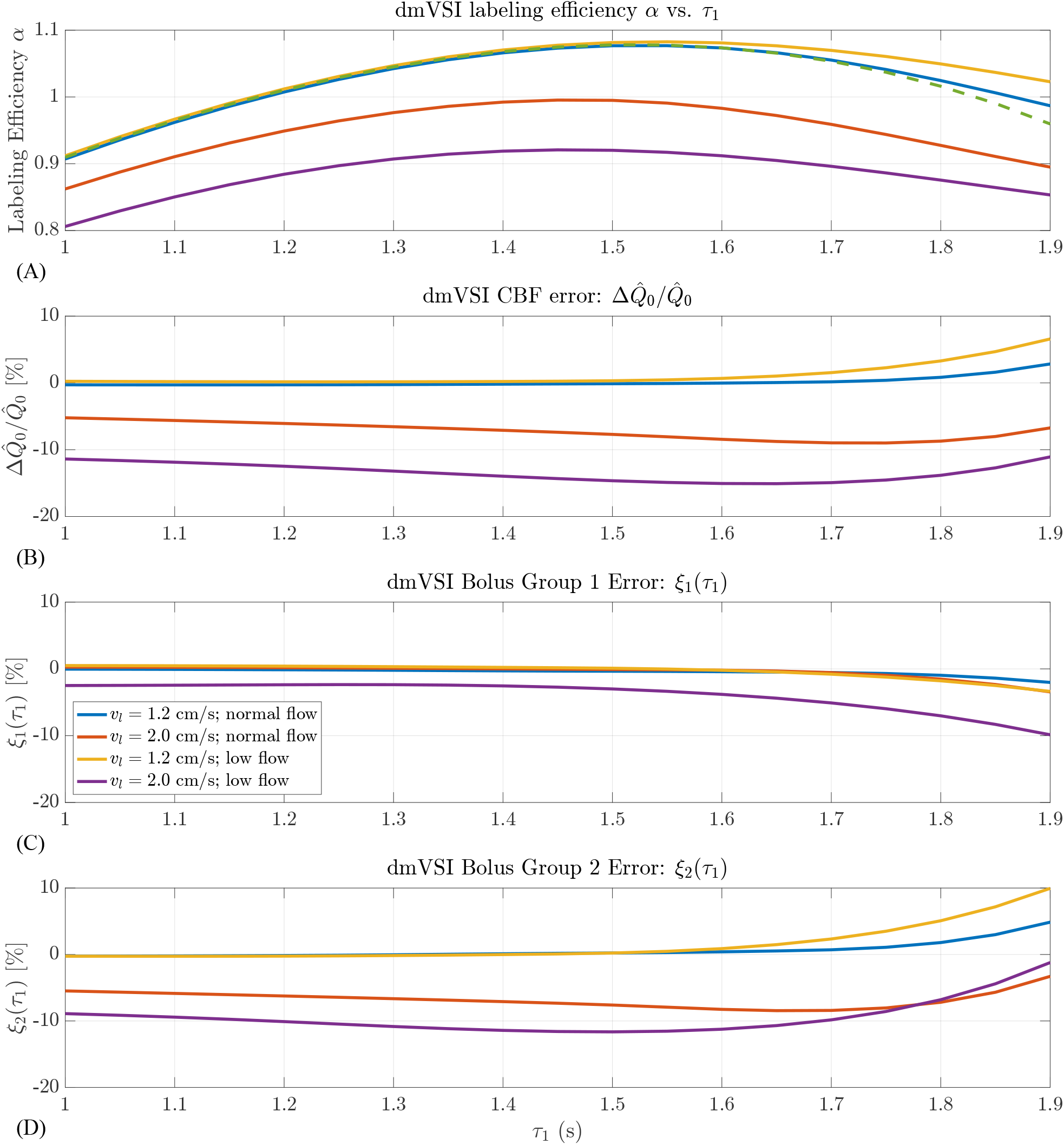
(A,B) dmVSI labeling efficiency and CBF estimation error as a function of *τ*_1_ for sinc FTVSI LCM (*v*_*l*_ = 1.2 or 2 cm/s as indicated in legend) and BIR8 VCM with *v*_*c*_ = 2, with nominal profiles The dashed green line in (A) indicates the labeling efficiency obtained with ideal rect profiles with *S*_0_ = *κ* = 1.0 and *β* = 0.88. (C,D) Error contributions from bolus groups 1 and 2. All errors are shown as percentages.

**FIGURE 5.**
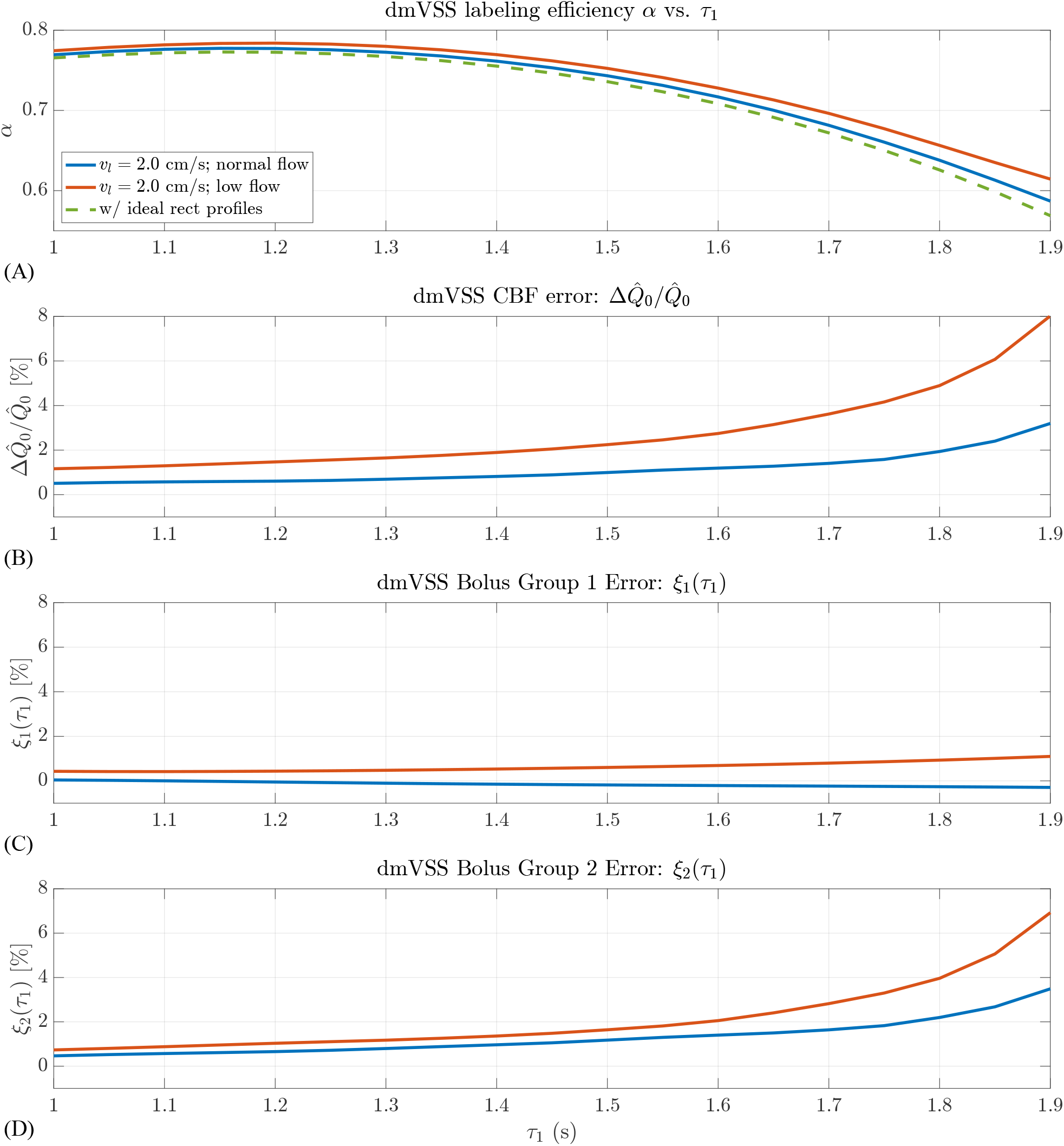
(A,B) dmVSS labeling efficiency and CBF estimation error as a function of *τ*_1_ for BIR8 LCM and VCM with *v*_*l*_ = *v*_*c*_ = 2, with nominal BIR8 profile. The dashed green line in (A) indicates the labeling efficiency obtained with ideal rect profiles with *S*_0_ = *κ* = *β* = 1.0. (C,D) Error contributions from bolus groups 1 and 2. All errors are shown as percentages.

It is also helpful to note that the fractional CBF quantification error can be written as

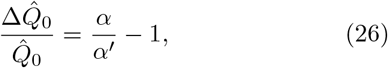

where *α* and *α*^*′*^ denote the actual and assumed labeling efficiencies, consistent with what was previously stated for single-module VSASL^7^.

### 2.6 Effective Bolus Width and Error

While the effective areas and related metrics, such as quantification error and labeling efficiency, can be readily calculated for arbitrary profiles, it is useful to write each effective area as the product of an amplitude term and an effective bolus width term^7^. For dmVSI, we have *A*_1,eff_ = 2*S*_0_*κ*^2^*βτ*_1,eff_ and *A*_2,eff_ = 2*S*_0_*κβτ*_2,eff_, where

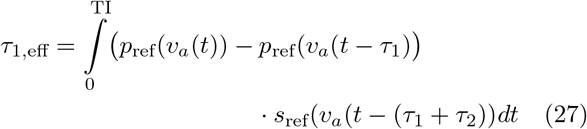

and

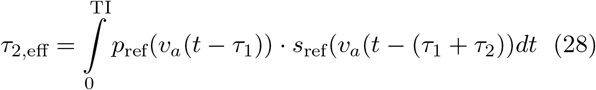

are the effective bolus widths. The fractional CBF estimation error for dmVSI can be written as

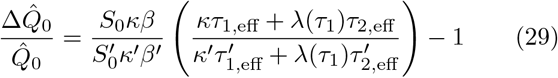

where the primed variables represent the assumed values associated with 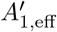 and 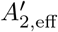. To isolate the estimation error due solely to bolus width errors, we may consider the case where the assumed amplitudes are equal to their actual values (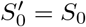, *κ*^*′*^ = *κ, β*^*′*^ = *β*), but the assumed bolus widths are equal to their nominal ideal values (i.e. 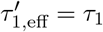 and 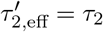, resulting in

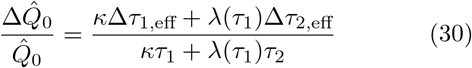

with bolus width errors defined as Δ*τ*_1,eff_ = *τ*_1,eff_ − *τ*_1_ and Δ*τ*_2,eff_ = *τ*_2,eff_ − *τ*_2_.

For dmVSS, *A*_2,eff_ = *S*_0_*κβτ*_2,eff_ where *τ*_2,eff_ was defined in Eqn. 28. Using the approximation from Eqn. 14, we can write *A*_1,eff_ ≈ *S*_0_*κ*^2^*β*(*τ*_[1,2],eff_ + (1 − *β*)*τ*_2,eff_) (see Section S.1) where

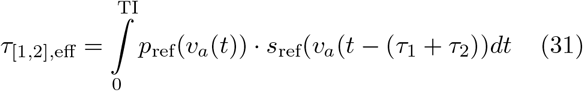

The fractional CBF estimation error for dmVSS can be written as

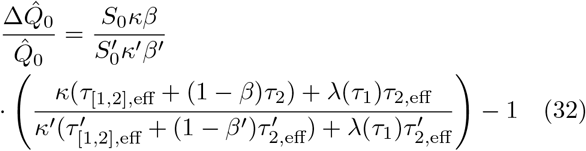

Isolating effects due solely to bolus width errors leads to

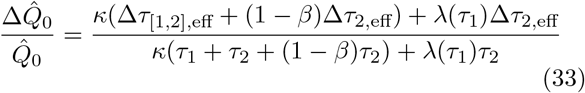

where Δ*τ*_[1,2],eff_ = *τ*_[1,2],eff_ − (*τ*_1_ + *τ*_2_).

### 2.7 Minimizing Bolus Width Error

While the effective bolus width errors can be calculated for arbitrary functions using Eqns. 27, 28 and 31, it is also useful to gain some theoretical insight into the conditions needed to obtain (*i*) Δ*τ*_1,eff_ = 0, (*ii*) Δ*τ*_2,eff_ = 0, and (*iii*) Δ*τ*_[1,2],eff_ = 0, which are sufficient for minimizing the CBF quantification errors described in Eqns. 30 and 33.

For case (*i*), if we assume that

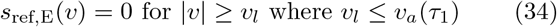

holds, as well as additional conditions min(*τ*_1_, *τ*_2_) > Δ*t*_l_ and *s*_ref_(*v*_b_) ≈ 1.0, then Eqn. 27 can be approximated as:

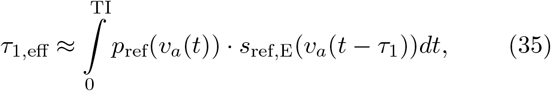

where *s*_ref,E_(*v*) = 1 − *p*_ref_(*v*) is an effective saturation function^7^. Because Eqn. 35 has the form of Eqn. 25 in Ref. [7] and Eqn. 34 is an extended version of condition W1 from that prior work (which we will denote as W1(*i*)), we may adopt the arguments from the prior work to state that the sufficient conditions to minimize the bolus width error for case (*i*) are: condition W1(*i*), an extended version W2(*i*): PLD ≥ Δ*t*_l_ − *τ*_2_ of condition W2 from the prior work, and the additional conditions.

Similarly, for cases (*ii*) and (*iii*), we note that the forms of Eqns. 28 and 31 follow that of Eqn. 25 in Ref. [7]. Sufficient conditions to minimize the bolus width errors for these cases are extended versions of condition W1: *s*_ref_(*v*) = 0 for |*v*| ≥ *v*_c_ where *v*_c_ ≤ *v*_a_(*τ*_2_) for condition W1(*ii*) and *v*_c_ ≤ *v*_a_(*τ*_1_ + *τ*_2_) for condition W1(*iii*); and an extended version W2(*ii, iii*): PLD ≥ Δ*t*_c_ of condition W2 that applies for both cases (*ii*) and (*iii*).

The expressions for the extended versions of Condition W1 are equivalent to: (i) *τ*_1_ ≥ Δ*t*_l_, (ii) *τ*_2_ ≥ Δ*t*_c_, and (iii) *τ*_1_ + *τ*_2_ ≥ Δ*t*_c_, where Δ*t*_l_ and Δ*t*_c_ are the LCM and VCM transit delays, respectively, described above in Sections 2.2 and 2.3. Since most dm-VSASL implementations use *τ*_1_ > *τ*_2_, the active design constraints are typically:

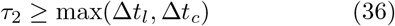

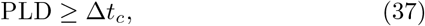

where we have taken into account the additional conditions for case (*i*). A summary of the conditions is provided in Table 2. With decreases in CBF, the transit delays are expected to increase^7^ and therefore the values of *τ*_2_ and PLD needed to meet the design constraints will also tend to increase.

**TABLE 2.** Summary of conditions for minimizing bolus width errors. In addition, we require min(*τ*_1_, *τ*_2_) > Δ*t*_*l*_ and *s*_ref_(*v*_*b*_) ≈ 1.0 for the approximation in Eqn 35 to hold. Since most implementations use *τ*_1_ > *τ*_2_ and typically Δ*t*_*l*_ ∼ Δ*t*_*c*_, the active design constraints are: *τ*_2_ ≥ max(Δ*t*_*l*_, Δ*t*_*c*_) and PLD ≥ Δ*t*_*c*_.

| Error Term | Bolus | Conditions |
| --- | --- | --- |
| $\Delta\tau_{1,eff}$ | dmVSI Bolus 1 | **W1(i):** $\tau_1 \geq \Delta t_l$<br>**W2(i):** $PLD \geq \Delta t_l - \tau_2$ |
| $\Delta\tau_{2,eff}$ | dmVSI or dmVSS Bolus 2 | **W1(ii):** $\tau_2 \geq \Delta t_c$<br>**W2(ii):** $PLD \geq \Delta t_c$ |
| $\Delta\tau_{[1,2],eff}$ | dmVSS Bolus 1 | **W1(iii):** $\tau_1 + \tau_2 \geq \Delta t_c$<br>**W2(iii):** $PLD \geq \Delta t_c$ |

Note that the conditions derived above for cases (*ii*) and (*iii*) assume the matched condition^7^ *s*_ref,E_(*v*) = *s*_ref_(*v*), which is applicable for dmVSS where the LCM1, LCM2, and VCM typically all use the same saturation module. For dmVSI, the matched condition holds for case (*i*) since LCM1 and LCM2 use the same inversion module, but typically does not hold for case (*ii*) due to differences in the LCM and VCM implementations, leading to the mismatch condition^7^ *s*_ref,E_(*v*) ≠ *s*_ref_(*v*) for the delivery of the second bolus *d*_a,2_(*t*). As discussed in Ref. [7] for single-module VSASL, this mismatch can lead to a CBF-dependent bolus width error that occurs when *v*_l_ = *v*_c_ and can be mitigated by setting *v*_l_ < *v*_c_. In Results, we show that the mismatch-related CBF estimation error for dmVSI (Δ*τ*_2,eff_ ≠ 0) can be similarly reduced with *v*_l_ < *v*_c_. Note that Fig. 6 in Ref. [7] provides a useful graphical interpretation of the mismatch error.

**FIGURE 6.**
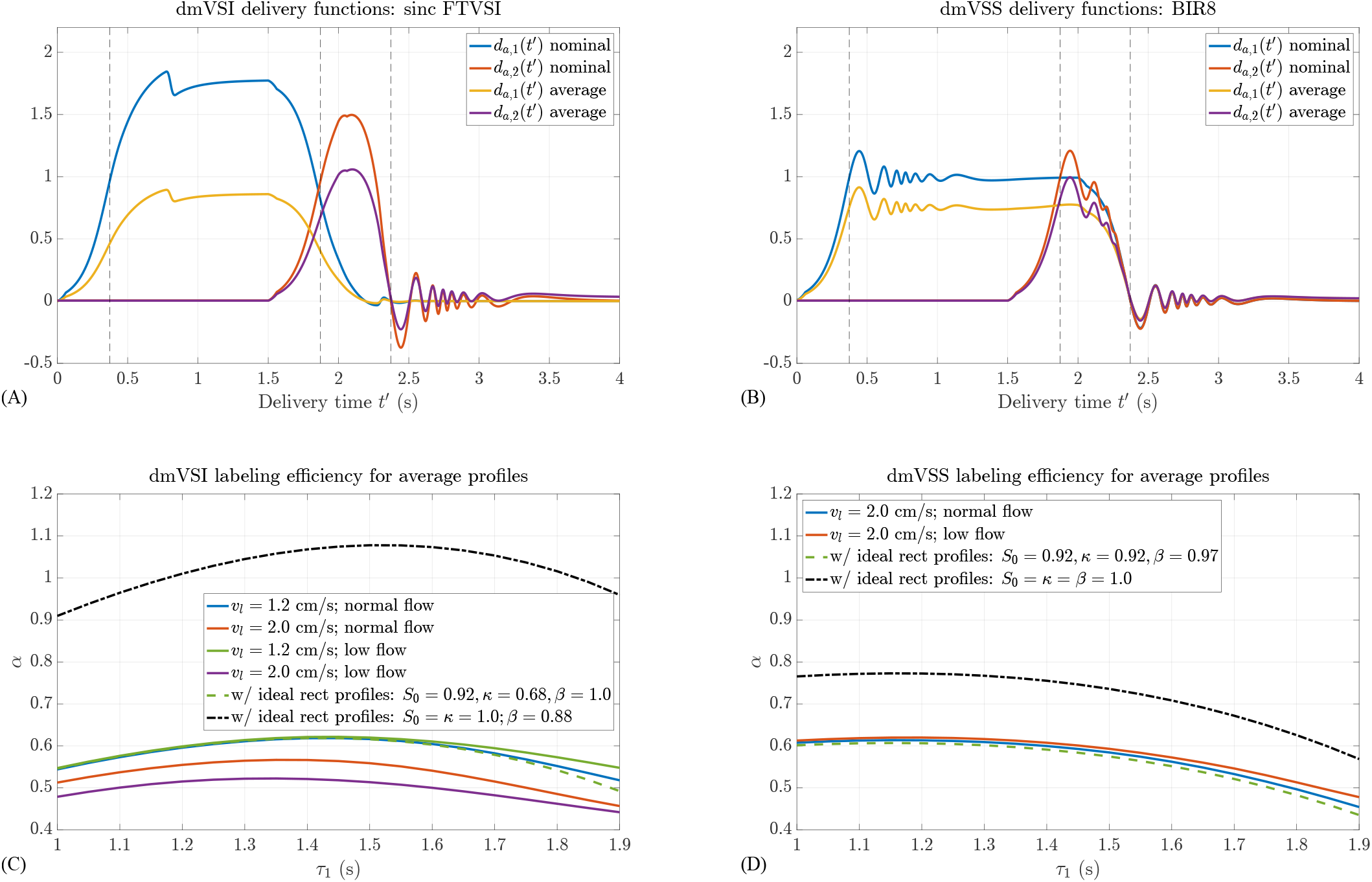
(A,B) dmVSI and dmVSS arterial delivery functions (yellow, purple; *v*_*l*_ = *v*_*c*_ = 2 cm/s, *v*_*b*_ = 0.1 cm/s) obtained using average profiles, with delivery functions for nominal profiles shown in blue and red for comparison (originally shown in Figures 2C and 3C). (C,D) dmVSI and dmVSS labeling efficiencies as a function of *τ*_1_ for (C) dmVSI with sinc FTVSI LCM (*v*_*l*_ = 1.2 or 2 cm/s as indicated in legend) and (D) dmVSS with BIR8 LCM with *v*_*l*_ = 2, with BIR8 VCM with *v*_*c*_ = 2 cm/s for both dmVSI and dmVSS. Labeling efficiencies with ideal rect profiles and scaling parameters from the average and nominal responses are shown by the green dashed lines and the black dash-dot lines, respectively.

### 2.8 Blood Volume Components

As noted in Ref. [7], the arterial delivery functions do not account for either (a) potential labeling of arterial blood in the velocity range [*v*_cap_, *v*_b_], where *v*_cap_ denotes capillary velocity, or (b) labeling of capillary and venous blood. Under the reasonable assumption that *p*_i_(*v*_cap_) ≈ 0, the contribution of the capillary blood volume term can be ignored^7^. Based on the similarity discussed above of Eqns. 28, 31, and 35 to Eqn. 25 in Ref. [7], the arguments from Appendix A.2 of the prior work can be adopted to show that the arterial blood volume terms are compensated for by negative adjustment terms in the bolus width computations, such that the arterial blood volume contributions can also be ignored. As a result, the prior treatment^7^ of local versus global model timing requirements for single-module VSASL also applies to dm-VSASL.

Adapting the arguments from Appendix A.6 of the prior work^7^ and assuming *τ*_1_ > *τ*_2_, minimization of the venous blood volume contribution requires that either (1) *τ*_2_ > Δ*t*_V,c_ or (2) *v*_v_(Δ*t*_V,c_ − *τ*_2_, *v*_cap_) is not much greater than *v*_cap_ where Δ*t*_V,c_ denotes the time for venous blood to accelerate from *v*_cap_ to *v*_c_ and *v*_v_(*t, v*_cap_) is the velocity at time *t* of a venous spin that starts with an initial velocity of *v*_cap_ at *t* = 0. When these conditions are not satisfied (i.e. for small values of *τ*_2_), Eqn. A.13 from Ref. [7] can be adapted to estimate the venous contribution.

## 3 METHODS

Bloch simulations and laminar flow integration were performed using code from the ISMRM 2022 VSASL Bloch Simulation Tutorial^8^. The BIR8 option with *v*_l_ = *v*_c_ = 2 cm/s was used to simulate the VSS module and the sinc FTVSI option was used to simulate the VSI module with *v*_l_ values of 1.2 and 2 cm/s (with velocity-dependent profiles shown in Figs. 2 and S1 of Ref. [7]). Key parameters were maximum *B*_1_ = 20 *µT*, maximum gradient 50 mT/m, maximum gradient slew rate 150 T/m/s, *T*_1_ and *T*_2_ set to ∞, and velocity span of 0 to 60 cm/s with a velocity increment of 0.001 cm/s. The profiles computed under these conditions are referred to as nominal profiles. The arterial acceleration model from Ref. [7] was used to calculate *v*_a_(*t*) assuming *v*_b_ = 0.1 cm/s for both normal (*Q*_0_ = 12.5 ml/s) and low flow conditions (*Q*_0_ = 7.7 ml/s).

Example equivalent arterial delivery functions and cumulative magnetization difference signals were then calculated assuming *T*_sat_ = 2.5 s, *τ*_1_ = 1.5 s, *τ*_2_ = 0.5 s, and *T*_1b_ = 1.66 s. For comparison, ideal rect VSI and VSS labeling functions with *v*_c_ = 2 cm/s and *κ* = *β* = *S*_0_ = 1.0 were also considered. Profiles were the same across LCM1 and LCM2, i.e. *l*_2_(*v*) = *l*_1_(*v*) and *c*_2_(*v*) = *c*_1_(*v*). We then varied *τ*_1_ from 1.0 s to 1.9 s (increment of 0.05 s) with *τ*_1_+*τ*_2_ = 2.0 s, and computed the delivery functions, magnetization difference signals, labeling efficiency, and CBF estimation error for each value of *τ*_1_.

Following the approach of Ref. [9], additional Bloch simulations were performed with *T*_1_ = 1660 ms, *T*_2_ = 150 ms, *B*_1_ scaled by a factor of 70% to 130% of its nominal value with an interval of 10%, and Δ*B*_0_ (off-resonance) varied from –200 Hz to 200 Hz with an increment of 50 Hz. Each of the resulting profiles was multiplied by a Gaussian weighting factor, computed from the product of two Gaussian distributions centered around mean values of *B*_1_ scale = 100% and Δ*B*_0_ = 0 Hz with standard deviations of 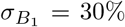 and 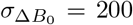 Hz, respectively. Average FTVSI and BIR8 profiles were then calculated as the sum of the weighted profiles. Labeling efficiency and CBF estimation error as a function of *τ*_1_ were then computed using the average profiles. Using the definitions from Section 2.2, the values of *κ* and *S*_0_ were set equal to the absolute values of the average labeling profiles of the LCM and VCM, respectively, at *v* = 0. The value of *β* was then determined by estimating the scaling factor needed to account for residual reductions in the steady-state value of the average passband function not accounted for by *κ*.

## 4 RESULTS

### 4.1 Nominal profiles

Example arterial delivery functions and cumulative magnetization difference signals computed using nominal profiles are shown in Fig. 2(C-F) for dmVSI (LCM: FTVSI with *v*_l_ = 2 or 1.2 cm/s; VCM: BIR8) and Fig. 3(C,D) for dmVSS (LCM and VCM: BIR8). As compared to the comparable waveforms obtained with ideal rect profiles, the delivery functions using the FTVSI and BIR8 waveforms exhibit smoothly varying leading and trailing edges. In addition, the value of the FTVSI cumulative magnetization difference at TI = *τ*_1_ + *τ*_2_ + Δ*t*_c_ is lower, reflecting the fact that *P*_a_ < 2.0 for the FTVSI passband function, as noted earlier in Section 2.5.

As shown in Fig. 4A, the dmVSI labeling efficiencies for both normal (blue) and low (yellow) flow with *v*_l_ = 1.2 cm/s are close to the ideal rect profile labeling efficiency (dashed green line) previously described in Section 2.5, and the corresponding CBF errors (blue and yellow lines Fig. 4B) are close to zero in the region of optimal *τ*_1_ ∼ [1.5, 1.6]. On the other hand, the labeling efficiencies with *v*_l_ = 2.0 cm/s are lower than that of the ideal rect profile, resulting in flow-dependent negative CBF errors (red and purple lines). These errors are largely due to the bolus group 2 error (red and purple lines in Fig. 4D), which reflects the mismatch described in Section 2.7. This error is consistent with the smaller area under the curve of *d*_a,2_(*t*) for *v*_l_ = 2 cm/s (Fig. 2C) as compared to that for *v*_l_ = 1.2 cm/s (Fig. 2E).

For the low flow condition with *v*_l_ = 2.0 cm/s, the bolus group 1 error (purple line in Fig. 4C) reflects the interaction between the edges of the *p*_ref_(*v*_a_(*t*)) and *p*_ref_(*v*_a_(*t* − *τ*_1_) terms in Eqn. 27, which vary more slowly for lower flow. For larger values of *τ*_1_, the corresponding values of *τ*_2_ = 2.0 − *τ*_1_ are too small to satisfy the condition stated in Eqn. 36, giving rise to the changes in the bolus group 2 errors. It is interesting to note that in this regime of small *τ*_2_ values, the errors associated with bolus groups 1 and 2 trend in different directions, such that the errors partially cancel out when computing the overall error. The negative trend for bolus group 1 reflects the interaction between the trailing edges of the LCM1 and VCM profiles, while the upward trend for bolus group 2 reflects the fact that for small *τ*_2_ there is less time for spins to decelerate such that the relevant portions of the leading edge of the LCM2 passband function occur at lower velocities and exhibit steeper slopes.

For dmVSS, the labeling efficiencies shown in Fig. 5A, for both normal and low flow with *v*_l_ = 2.0 cm/s, closely follow the ideal rect profile labeling efficiency (dashed green line) described in Section 2.5. As a result, the corresponding CBF errors (Fig. 5B) are relatively small (< 2%) in the region of optimal *τ*_1_ ∼ [1.0, 1.2], reflecting the inherent matching of the LCM and VCM functions described in Section 2.7. For larger values of *τ*_1_ where the condition stated in Eqn. 36 is violated, the increase in the bolus group 2 error (Fig. 5D), drives the overall increase in error.

### 4.2 Average profiles

Delivery functions using average profiles (i.e sum of weighted profiles over range of *B*_1_ and Δ*B*_0_ values as described in Methods) are shown by the yellow and purple curves in panels A and B of Fig. 6 for dmVSI and dmVSS, respectively, with the corresponding delivery functions using nominal profiles (originally depicted in Figs. 2C and 3C) shown by the blue and red curves for comparison. With the average profiles, the amplitudes of the dmVSI delivery functions are considerably smaller than those for the nominal profiles. These are consistent with the predicted amplitudes of 2*S*_0_*κ*^2^*β* and 2*S*_0_*κβ* for *d*_a,1_(*t*) and *d*_a,2_(*t*), respectively, where *S*_0_ = 0.92, *κ* = 0.68, and *β* = 1.0 for the average profiles as compared to *S*_0_ = *κ* = 1.0, and *β* = 0.88 for the nominal profiles. Note that for the average profiles the amplitude of *d*_a,1_(*t*) is more attenuated than that of *d*_a,2_(*t*), reflecting the difference in their respective dependence on *κ* (i.e. ∼ *κ*^2^ vs. ∼ *κ*).

For the dmVSS average profiles, the reduction in the delivery function amplitudes are consistent with the predicted amplitudes of ∼ *S*_0_*κ*^2^*β* (approximation for *β* close to 1.0) and *S*_0_*κβ* for *d*_a,1_(*t*) and *d*_a,2_(*t*), respectively, where *S*_0_ = 0.92, *κ* = 0.92, and *β* = 0.97 for the average profiles as compared to *S*_0_ = *κ* = *β* = 1.0 for the nominal profiles. Similar to what was observed for dmVSI, the amplitude of *d*_a,1_(*t*) shows greater attenuation than that of *d*_a,2_(*t*) due to the *κ*^2^ dependence.

Labeling efficiencies (both normal and low flow conditions) for the average profiles are shown in panels C and D of Fig. 6 for dmVSI and dmVSS, respectively, with the corresponding labeling efficiencies using (i) the ideal rect profiles (with the *S*_0_, *κ*, and *β* values of the average profiles) shown for reference and (ii) the ideal rect profiles with the scaling parameters from the nominal profiles (originally shown in Figs. 4A and 5A) shown for comparison. For dmVSI, the *v*_l_ = 1.2 cm/s curves show good agreement with the ideal rect reference curve, whereas the *v*_l_ = 2 cm/s show a flow-dependent negative bias, similar to what was observed for the nominal profiles shown in Figs. 4A. Consistent with the markedly lower dmVSI delivery function amplitudes shown in Fig. 6A, the dmVSI labeling efficiencies are greatly attenuated for the average profiles as compared to the nominal profiles.

The dmVSS labeling efficiencies exhibit a moderate degree of attenuation with the average profiles, reflecting the greater robustness of the BIR8 profile to *B*_0_ and *B*_1_ inhomogeneities. The maximum dmVSI and dmVSS labeling efficiencies assuming average profiles are both approximately 0.6, in marked contrast to the difference in dmVSI and dmVSS maximum labeling efficiencies obtained for the nominal profiles.

## 5 DISCUSSION AND CONCLUSIONS

We have presented a generalized signal model for dm-VSASL that can integrate arbitrary saturation and inversion profiles. In addition, we have shown how the framework of passband and saturation reference functions originally developed for the analysis of single-module VSASL^7^ can facilitate the analysis and interpretation of the signals, including the derivation of concise expressions for labeling efficiency and CBF quantification error.

For the case of scaled ideal rect profiles, the signal expressions match those of the prior work^2^, with the addition of a minor correction term for dmVSS that addresses a small error in the prior work. To facilitate comparison with the prior work, we defined scaling terms *κ* and *β*, where *κ* denotes the attenuation that is common to *l*(*v*) and *c*(*v*) at *v* = 0 and *β* accounts for additional scaling of the passband function that is not accounted for by *κ*. This definition supports the derivation of labeling efficiency expressions (Eqns. 24 and 25) for scaled ideal rect profiles that can be used to represent the expected labeling efficiency in the absence of bolus width errors (e.g. green dashed lines in Figs. 4A, 5A, and 6(C,D)). It is important to note, however, that our definition of *κ* and *β* differs from the prior work^2^, in which it was assumed that *κ* accounted only for the effects of *T*_2_ decay and *β* accounted for all other effects. While the product *κβ* is the same across definitions, the definition presented here is preferred for future work as it is grounded in the generalized signal model and thereby facilitates a rigorous analysis and interpretation of dm-VSASL labeling efficiency and CBF quantification error.

When using realistic Bloch-simulated profiles, we demonstrated that flow-dependent CBF quantification errors can occur for dmVSI due to the mismatch in the profiles of the LCM and VCM functions, similar to those observed for single-module VSI^7^. This error is primarily due to the mismatch of leading and trailing edges in the second bolus and can be mitigated by selecting a lower labeling cutoff velocity *v*_l_ < *v*_c_, consistent with the findings for single-module VSI^7^. For small values of *τ*_2_, we observed opposing negative and positive trends in the bolus group 1 and 2 errors that partly canceled out in the overall error. Further work to identify approaches for maximizing the cancelation could be of interest.

In this work, we assumed that LCM1 and LCM2 used the same labeling profiles, with matched VCM for dmVSS and mismatched VCM for dmVSI. In the prior modeling of single-module VSASL^7^, it was noted that there may be cases where where the mismatch between the LCM and VCM profiles could be of interest, for example to characterize the volume of arterial blood in various velocity ranges. For dm-VSASL, there is an even wider space of potential mismatches to consider due to the presence of three modules (LCM1, LCM2, and VCM), and further work to explore this space would be of interest.

Conditions for minimizing bolus width errors were derived under the assumption of matched profiles. For most implementations, where *τ*_1_ > *τ*_2_, the active requirements are that *τ*_2_ is greater than both the LCM and VCM transit delays and that the PLD is greater than the VCM transit delay. As discussed in Ref. [7], these transit delays are likely to exhibit a dependence on voxel size, with preliminary model-based estimates on the order of 100 to 200 ms when *v*_c_ = *v*_l_ = 2 cm/s. While further experimental work is needed to refine the estimates, researchers wishing to minimize bolus width errors may want to consider a minimum value of at least 100 to 200 ms for both PLD and *τ*_2_ as an interim design rule.

Consistent with prior work we found that the labeling efficiency of dmVSI was higher than that of dmVSS under the assumption of nominal profiles (with *T*_1_ = *T*_2_ = ∞ and nominal *B*_0_ and *B*_1_ values). However, when taking into account *B*_0_ and *B*_1_ inhomogeneities, the labeling efficiencies were found to be comparable, reflecting the the greater robustness of BIR8 dmVSS to the inhomogeneities^9^. While this is roughly consistent with prior in vivo findings^9^ of comparably high ASL signal levels with BIR8 dmVSS and sinc FTVSI VSI (which were found to be about 5% lower than sinc FTVSI dmVSI ASL signal levels^2^), further work is needed to more accurately assess the empirical effect of the inhomogeneities in vivo (see also Section S.2).

As noted in this paper, dmVSS has the advantage of inherent matching of LCM and VCM profiles, resulting in inherently small CBF quantification errors when using *v*_l_ = *v*_c_. On the other hand, the inversion of static tissue in dmVSI (or a dm-VSASL variant that uses VSSI pulses) provides inherent background suppression that can lead to improved temporal SNR even without additional background suppression pulses^6^. Due to the label/control condition switching in the second module, dmVSI also offers a more balanced distribution of motion-sensitive gradients, which can reduce artifacts from sources such as eddy currents and diffusion attenuation^2^. The effect of these sources on the dm-VSASL signal can be examined with the generalized signal model through the inclusion of saturation and inversion profiles that reflect the presence of the confounding factors, and further work in this area would be of interest.

Consistent with the prior work we have assumed that the *τ*_1_ + *τ*_2_ < *BD*_max_, where *BD*_max_ denotes the maximum available physical bolus width and depends on the spatial coverage of the RF transmit coil and the mean velocity of arterial spins^1,2^. This is equivalent to neglecting the contribution of arterial spins that enter the coverage of the RF transmit coil after the first LCM. Extensions of the current model to model this contribution would be of interest and would most likely entail introducing a joint dependence on velocity and space into the passband function that would allow for an estimation of the physical bolus width.

In this paper we have focused on dm-VSASL, which is currently the primary implementation of multi-module VSASL, as prior work indicates that there is unlikely to be an advantage to using more than two modules^1^. The challenges of using more than two modules include further decreases in labeling efficiency with the addition of each module and potential concerns with specific absorption rate, especially at higher magnetic field strengths^1,2^. Nevertheless, if VSASL with more than two modules becomes of interest for future work, the approach described in the Appendix can be readily extended to handle additional modules.

In conclusion, by facilitating the use of realistic saturation and inversion profiles, the proposed signal model can provide a more accurate assessment of the performance (e.g. labeling efficiency) of dm-VSASL implementations and improve the quantification of dm-VSASL CBF measures.

## 6 DATA AVAILABILITY STATEMENT

Analysis code and files to generate the figures and results presented in this paper will be made available upon publication through the Open Science Framework DOI:10.17605/OSF.IO/WKM54.

## SUPPORTING INFORMATION

The following supporting information is available as part of the online article:

**Figure S1**. Example dmVSI and pCASL normalized ASL images Δ*M*_z_*/M*_0b_ in percentage units.

How to cite this article

Liu TT, Chen C, Guo J, Wong EC, and Bolar DS (2024), A Generalized Signal Model for Dual-Module Velocity-Selective Arterial Spin Labeling, *Magn. Reson. Med*.,.

## APPENDIX

Define 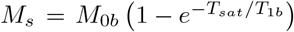 and the longitudinal recovery function for the blood signal as

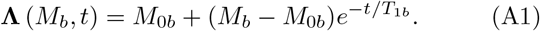

Note that **Λ** (*M*_*b*_, *t*) is a general-purpose function that returns the longitudinal magnetization given an initial condition *M*_*b*_ and relaxation time *t*, whereas the longitudinal relaxation weighting functions *L*_1_(*t*) and *L*_2_(*t*) defined in Section 2.1 can be obtained from the expressions provided in the latter part of this Appendix. We use the notation 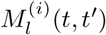 and 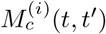 to denote the label and control, respectively, magnetization at experimental time *t* for blood that is delivered at *t*^*′*^ to the boundary velocity *v*_b_, where the subscript (*i*) indicates the *i*th step in the processing beginning with LCM1 as the first step (*i* = 1). With the application of the LCM1 at *t* = 0, we have

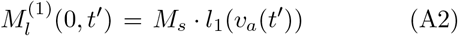

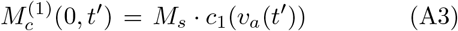

Over the interval *t* ∈ [0, *τ*_1_) the longitudinal magnetization is

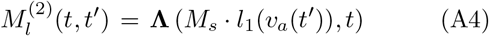

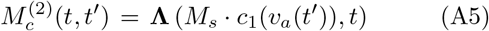

At the point of the application of LCM2 at *t* = *τ*_1_, we have

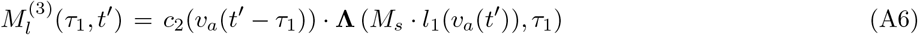

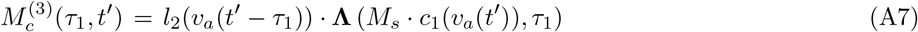

for dmVSI and

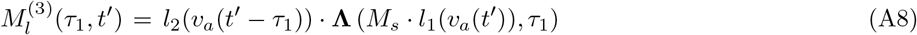

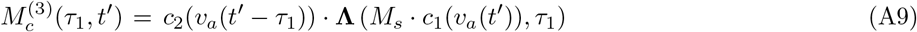

for dmVSS. The magnetization difference at this point is

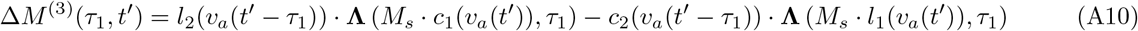

for dmVSI and

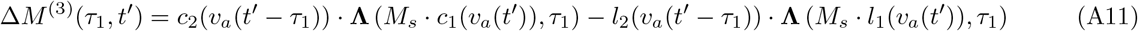

for dmVSS. After expanding and reorganizing terms, these expressions may be rewritten as

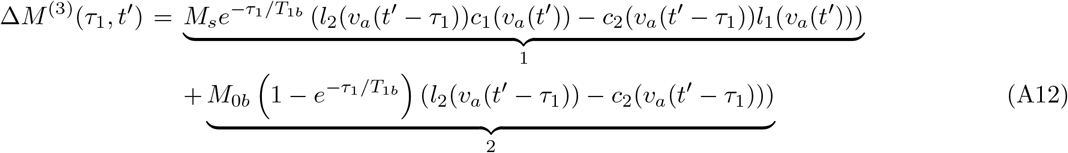

for dmVSI and

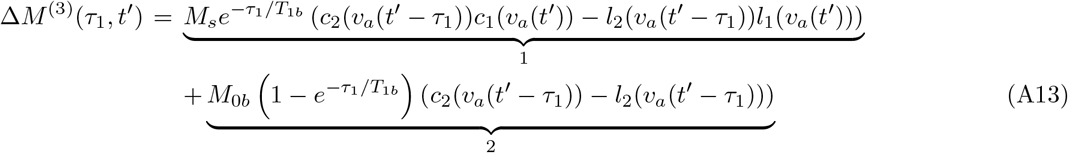

for dmVSS, where term 1 reflects the effects of both LCM1 and LCM2 and term 2 reflects the effect of only LCM2.

After the application of LCM2, both the control and label paths go through the same longitudinal relaxation and VCM, so that the magnetization difference for time *t* > *τ*_1_ + *τ*_2_ may be written as

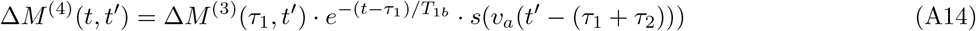

To obtain the cumulative magnetization difference at time *t*, we integrate over all spins that were delivered over the interval *t*^*′*^ ∈ [0, *t*], resulting in

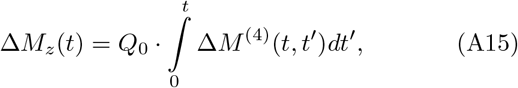

which can be expanded and rewritten as the equations presented in Section 2.

