## Supplementary Information for "A Generalized Signal Model for Dual-Module Velocity-Selective Arterial Spin Labeling"

*Running Head:* Dual-Module VSASL Signal Model


*Address correspondence to:*

Thomas Liu, UCSD Center for fMRI, 9500 Gilman Drive MC 0677, La Jolla, CA 92093



Supporting Information submitted to Magnetic Resonance in Medicine

### S.1 Derivation of Equation 31

Using the approximation

$$\Pi_1(t) \approx \kappa^2\beta \cdot \big(p_{\mathrm{ref}}(v_a(t)) + (1-\beta)p_{\mathrm{ref}}(v_a(t-\tau_1))\big) \tag{S.1}$$

stated in Equation 14equation.2.14 of the text, we can write

$$\begin{aligned}
A_{1,\mathrm{eff}} &= \frac{1}{Q_0}\int_0^{\mathrm{TI}} d_{a,1}(t)dt && \text{(S.2)}\\
&\approx S_0\kappa^2\beta \int_0^{\mathrm{TI}} \big(p_{\mathrm{ref}}(v_a(t)) + (1-\beta)p_{\mathrm{ref}}(v_a(t-\tau_1))\big) \cdot s_{\mathrm{ref}}(v_a(t-(\tau_1+\tau_2))dt && \text{(S.3)}\\
&= S_0\kappa^2\beta \left(\int_0^{\mathrm{TI}} p_{\mathrm{ref}}(v_a(t)) \cdot s_{\mathrm{ref}}(v_a(t-(\tau_1+\tau_2))dt + \right.\\
&\left. (1-\beta)\int_0^{\mathrm{TI}} p_{\mathrm{ref}}(v_a(t-\tau_1)) \cdot s_{\mathrm{ref}}(v_a(t-(\tau_1+\tau_2))dt\right) && \text{(S.4)}\\
&= S_0\kappa^2\beta \left(\tau_{[1,2],\mathrm{eff}} + (1-\beta)\tau_{2,\mathrm{eff}}\right) && \text{(S.5)}
\end{aligned}$$

where

$$\tau_{[1,2],\mathrm{eff}} = \int_0^{\mathrm{TI}} p_{\mathrm{ref}}(v_a(t)) \cdot s_{\mathrm{ref}}(v_a(t-(\tau_1+\tau_2))dt \tag{S.6}$$

and

$$\tau_{2,\mathrm{eff}} = \int_0^{\mathrm{TI}} p_{\mathrm{ref}}(v_a(t-\tau_1)) \cdot s_{\mathrm{ref}}(v_a(t-(\tau_1+\tau_2))dt. \tag{S.7}$$

### S.2 Example ASL Images

Figure S.1 shows dmVSI normalized ASL images ($\Delta M_z/M_{0b}$ in percentage units) with comparison pCASL images for a representative subject, with dmVSI parameters: $\tau_1 = 1.6$ s, $\tau_2 = 0.4$ s, $T_{sat} = 2.0$ s, PLD = 0.1 s, sinc FTVSI LCM with phase-cycling and $v_l = 2.0$ cm/s, BIR-8 VCM with $v_c = 2.0$ cm/s, no additional background suppression pulses (inherent background suppression provided by dmVSI (Guo, 2024)); pCASL parameters: $\tau = 1.8$ s, PLD = 2.0 s, 4 background suppression pulses; and imaging parameters (same across dmVSI and pCASL): single-shot 3D gradient and spin echo (GRASE) readout, voxel size=3.5 ×3.5 ×5 $mm^3$, FOV=224 ×224 ×160 $mm^3$, 32 slices, TE = 12 ms, EPI factor = 15, turbo spin echo factor = 16, readout duration for each EPI module = 8 ms, total readout duration = 220 ms, bandwidth = 2604 Hz/pixel, TR = 4600 ms, parallel acquisition technique acceleration = 3 × 2 in PExSlice [GRAPPA], 28 ASL measurements, one $M_0$ measurement.

The slightly higher normalized ASL values for the dmVSI images as compared to the pCASL images are roughly consistent with predicted percentage values of

$$100 \cdot \frac{\Delta M_z}{M_{0b}} = 2 \cdot 100 \cdot Q_0 \cdot \alpha_{\text{dmVSI}} \cdot (\tau_1 + \tau_2) \cdot \left(1 - e^{-T_{sat}/T_{1b}}\right) \cdot e^{-(\tau_1+\tau_2+\text{PLD})/T_{1b}} \sim 0.43 \tag{S.8}$$

for dmVSI where $\alpha_{\text{dmVSI}} = 0.54$ (using the average profiles and normal flow acceleration model results shown in Figure 6 of the main text) and we have assumed an average CBF of $Q_0 \sim 60\, ml/(100\, g\text{-}min) \sim 0.01\, s^{-1}$ so that $100 \cdot Q_0 \sim 1.0\, s^{-1}$; and

$$100 \cdot \frac{\Delta M_z}{M_{0b}} = 2 \cdot 100 \cdot Q_0 \cdot \alpha_{\text{pCASL}} \cdot T_{1b} \cdot \left(1 - e^{-\tau/T_{1b}}\right) \cdot e^{-\text{PLD}/T_{1b}} \sim 0.40 \tag{S.9}$$

for pCASL, where we have assumed a labeling efficiency $\alpha_{\text{pCASL}} = 0.60$ that includes the attenuation effects of the four background suppression pulses (Alsop et al., 2015).

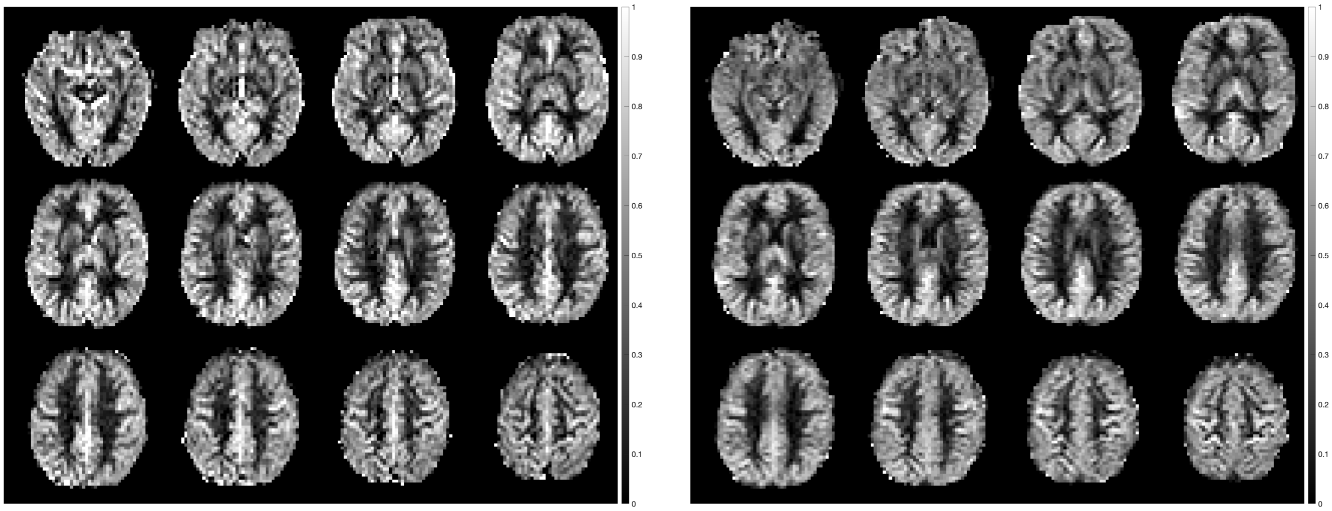

(a) dmVSI

(b) pCASL

Figure S.1: Example dmVSI and pCASL normalized ASL images $\Delta M_z/M_{0b}$ in percentage units.
